# Carer strain in post-stroke emotionalism: an initial evaluation

**DOI:** 10.1101/2023.11.06.23298191

**Authors:** Niall M Broomfield, Matthew Walters, Robert M West

**Affiliations:** Department of Clinical Psychology and Psychological Therapies, Norwich Medical School, University of East Anglia, NR4 7TJ UK; Institute of Cardiovascular and Medical Sciences, University of Glasgow, G12 8QQ UK; Leeds Institute of Health Sciences, University of Leeds, LS2 9JT UK

**Keywords:** Stroke, Emotionalism, Mental health, Carer strain, Caregiver burden

## Abstract

**Background and Purpose:** Post-stroke emotionalism is a common, under-researched neurologic symptom of stroke, characterised by frequent crying episodes not under usual social control. Currently there are no data on carer strain in the context of emotionalism after stroke.

**Aims:** To explore the degree of carer strain in carers of individuals with diagnosed PSE compared to carers of individuals with stroke but no PSE. To examine whether carer strain varies with particular characteristics of the cared for individual: patient age, sex, social deprivation, stroke type, functional status, mood status.

**Methods:** 102 informants of people with stroke completed the Modified Carer Strain Index at 6-months as part of the TEARS longitudinal cohort study between October 1^st^ 2015 and September 30^th^ 2018. Stroke survivor diagnostic status was determined using Testing Emotionalism After Recent Stroke-Diagnostic Interview based on published, widely accepted diagnostic criteria of emotionalism.

**Results:** There was little evidence of association between carer strain and sex, age, deprivation level or stroke type of the cared for individual. There was strong evidence that carer strain associated with both increased functional dependence post-stroke, and presence of post-stroke emotionalism.

**Conclusions:** Even after accounting for increased functional dependence, our study data indicates that caring in a PSE context may significantly increase carer strain, comparable to a six-point reduction on Barthel Index.

## BACKGROUND

Post-stroke emotionalism (PSE) is a widely acknowledged, clinically prevalent neurological stroke sequela promoting distress, disrupted stroke rehabilitation and eroded social functioning and quality of life. PSE is characterised by frequent, sudden onset crying episodes (occasionally laughter), not under usual social control which represent a change from pre-stroke functioning.^1–3^

Surprisingly, despite the prevalence and nature of PSE and its psychological associations ^4–8^, little is known regarding the impact of caring for people living with PSE.

Colamonico and colleagues progressed an online survey which included carers of people with and without emotionalism following a range of precipitating neurologic diseases, including stroke.^9^ Three self-report measures were completed by the carers: the Center for Epidemiologic Studies Depression Scale 10-item (CESD-10)^10^, the Screen for Caregiver Burden (SCB)^11^ and the Work Productivity and Activity Impairment questionnaire (WPAI).^12,13^ Interestingly, those caring for people with emotionalism showed equivalent levels of depression to carers of people without emotionalism. Whereas, amongst the emotionalism carers, there were more frequent burdensome events, higher distress levels, greater disruption to work productivity and work impairment, including missed time at work, and more caregiver burden overall.^9^

These are important data. They suggest increased carer burden and occupational dysfunction linked specifically to emotionalism, over and above the impact of caring in the context of the index neurologic disease without concomitant emotionalism.

Notably however carers in the Colamonico survey were from a mixed neurologic disease sample. Under one-sixth of the carer sample were caring specifically in the context of stroke (N = 59) and data specific to caring in the context of stroke emotionalism are not separately reported. ^9^ Thus, the specific psychological impact of caring for people with emotionalism after stroke is not known, and we could find no other studies which have explored this topic directly.

Our primary aim was to address this knowledge gap, via an analysis of Modified Caregiver Strain Index (MCSI)^14^ data, collected from carers of individuals with or without diagnosed PSE 6-months post index stroke derived from the TEARS (Testing Emotionalism After recent Stroke; NRS Stroke Research Network ID 18980) longitudinal cohort study.^15^

## AIMS

As the TEARS data set contains MCSI scores from carers of individuals with stroke and PSE but also stroke and no PSE, the study had two aims:

[i] To explore the degree of carer strain in carers of individuals with diagnosed PSE and to compare this to the degree of carer strain in carers of individuals with stroke but no diagnosis of PSE
[ii] To examine whether carer strain varies with particular characteristics of the cared for individual: patient age, sex, social deprivation, stroke type, functional status, mood status (depression, anxiety).
[iii] To quantify the impact of PSE on carer strain after accounting for other factors.

## METHODS

### Sample Size

The TEARS cohort study recruited N = 277 participants. Of these, N=102 had informants at 6-month follow up reporting carer strain data. Within these, at sixty-six were shown not to have PSE at 6-months, sixteen to have PSE at 6-months and twenty not to have PSE status determined. To compare a normally distributed outcome measure between the sixty-six non-PSE and sixteen PSE carers, there would be 81% power to detect a standardised measure of 0.8 and 94% power to detect a standardised measure of 1.0. These differences can be compared to the range of MSCI which is 0-26. Thus, there was sufficient power to detect clinical differences in the continuous outcome provided it was sufficiently close to normal in distribution.

### Participants

Participants were recruited into TEARS from nine Scottish hospital acute stroke units within two weeks of sustaining stroke (https://www.stroke.org.uk/research/understanding-difficulty-controlling-emotions-after-stroke; full protocol available from NB) between October 1^st^ 2015 and September 30^th^ 2018. For each participant, an informant (spouse or closest relative) was recruited.

All stroke participants were male or non-pregnant female, ≥18 years of age, with clinical diagnosis of first ever or repeat ischaemic or haemorrhagic stroke. We excluded on the basis of subarachnoid haemorrhage, other extra-axial bleeds, Transient Ischemic Attack, severe concurrent medical conditions, severe distressing behaviours precluding participation, aphasia (score < 25 on Frenchay Aphasia Screening Test^16^), life expectancy ≤ 3 months and/or lack of spoken English. We did not gather data on informant respondent characteristics, other than confirming next of kin or equivalent to the stroke participant.

### Measures and Procedure

TEARS had a priori ethical approval from the Scotland A Research Ethics Committee (IRAS Reference 157483). All participants gave written informed consent, including those informant participants whose data are reported here.

Findings are based on data (all measures) gathered face-to-face at the 6-months assessment point by pre-trained stroke research nurses based on the stroke units.

Carer strain was determined using the MCSI.^14^ The scale was developed for use with family carers based on the original Caregiver Strain Index version.^17^ MCSI is self-report, contains thirteen items including prompting examples across a range of pertinent strain domains (physical, personal, psychological, financial, social) with item responses captured as follows: ‘Yes, on a regular basis’ (scored 2), ‘Yes, sometimes’ (scored 1) or ‘No’(scored 0). This gives a possible total score ranging from 0 (no carer strain) to 26 (highest carer strain).

MCSI has acceptable internal (alpha=.90) and test-retest reliability (alpha=.86) basing on the Thornton and Travis sample^14^, and has been used previously to screen carer burden in the context of neurologic disease^18,19^, including stroke.^20^

Diagnosis of PSE status was reached using Testing Emotionalism After Recent Stroke-Diagnostic Interview (TEARS-IV)^15^, a semi-structured diagnostic interview based on published, widely accepted diagnostic criteria of emotionalism.^1,4,5,21^

We classified stroke using the Oxford Classification System^22^ and used Hospital Anxiety and Depression Scale (HADS)^23^ to measure anxiety and depression, Barthel Activities of Daily Living Index^24^ for functional outcome and computed deprivation level basing on Scottish Government Index of Multiple Deprivation (SIMD), scaled by 1000 ranks.^25^

## Statistical Analysis

All inferential analyses were conducted using R-Software, version 4.2.3.^26^ Initial cross tabulation and follow up statistics characterised the sub-sample from whom carer strain data were collected. Cross tabulation was used to explore association of carer strain (MCSI) with stroke classification recorded at baseline, and PSE status (Yes/No on TEARS-IV), anxiety (HADS-A), depression (HADS-D) and Barthel Activities of Daily Living Index measured at 6-months follow up when MCSI was also measured.

On initial inspection, MCSI carer strain data had a skewed distribution. To improve variance stabilisation, we used the square-root of MCSI, hereafter sqrt.MCSI, for modelling.

Univariate analyses were undertaken, regressing sqrt.MCSI on each covariate separately to identify covariates with the strongest association with sqrt.MCSI. Then a suitable regression model was fitted. In particular, the interest was focussed on PSE which was considered to lead to greater anxiety and depression. Thus the model for sqrt.MCSI was regressed upon PSE status and other covariates but not HADS-A nor HADS-D. The final model was selected to be a parsimonious fit for which all regression coefficients were statistically significant at the 5% level.

## RESULTS

### Participants

Characteristics of the final sample are shown in Table 1.

**Table 1:**
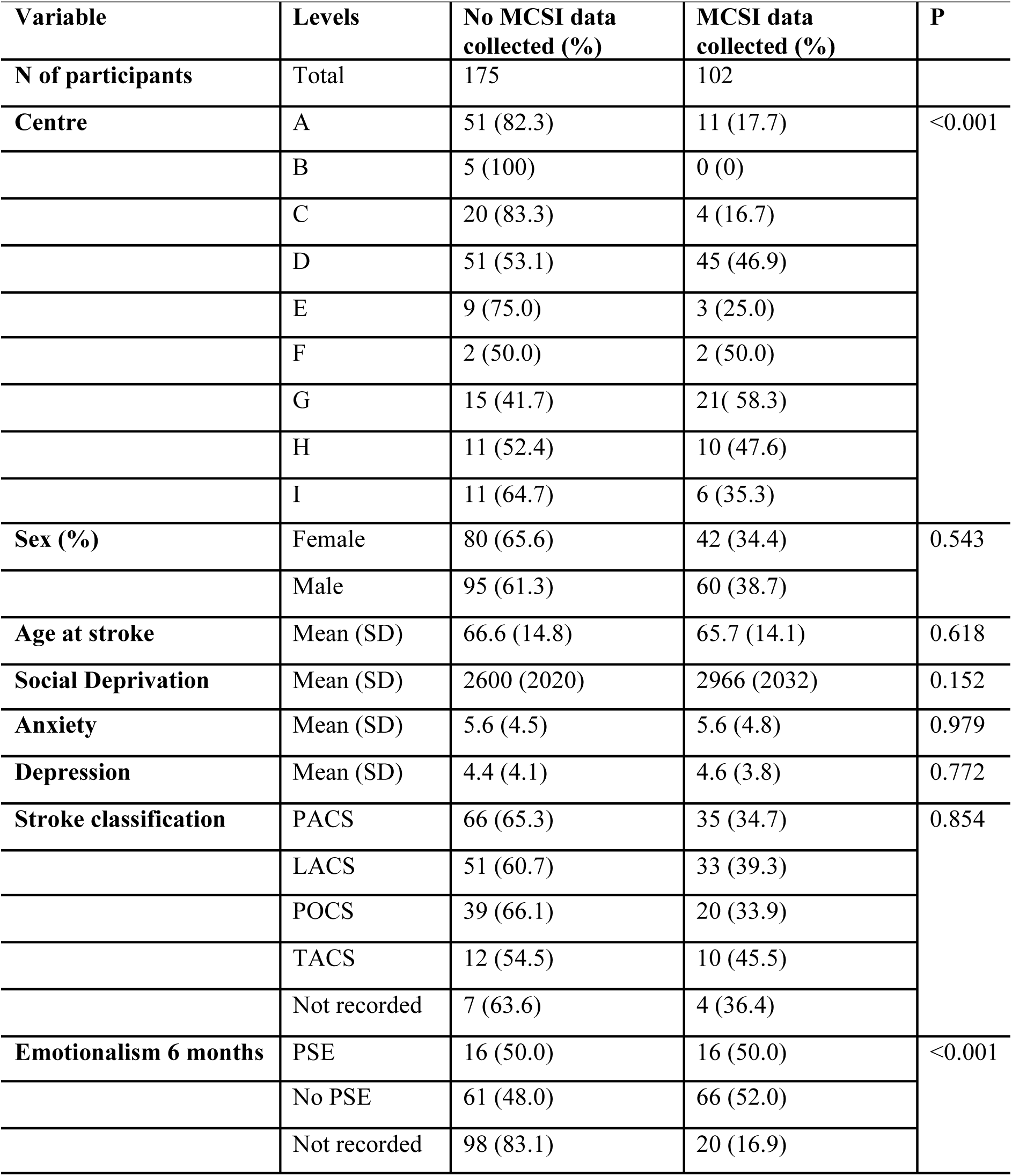
Participant characteristics, cross-classified by collection of caregiver strain on MCSI.

| Variable | Levels | No MCSI data collected (%) | MCSI data collected (%) | P |
| --- | --- | --- | --- | --- |
| **N of participants** | Total | 175 | 102 |  |
| **Centre** | A | 51 (82.3) | 11 (17.7) | <0.001 |
|  | B | 5 (100) | 0 (0) |  |
|  | C | 20 (83.3) | 4 (16.7) |  |
|  | D | 51 (53.1) | 45 (46.9) |  |
|  | E | 9 (75.0) | 3 (25.0) |  |
|  | F | 2 (50.0) | 2 (50.0) |  |
|  | G | 15 (41.7) | 21 (58.3) |  |
|  | H | 11 (52.4) | 10 (47.6) |  |
|  | I | 11 (64.7) | 6 (35.3) |  |
| **Sex (%)** | Female | 80 (65.6) | 42 (34.4) | 0.543 |
|  | Male | 95 (61.3) | 60 (38.7) |  |
| **Age at stroke** | Mean (SD) | 66.6 (14.8) | 65.7 (14.1) | 0.618 |
| **Social Deprivation** | Mean (SD) | 2600 (2020) | 2966 (2032) | 0.152 |
| **Anxiety** | Mean (SD) | 5.6 (4.5) | 5.6 (4.8) | 0.979 |
| **Depression** | Mean (SD) | 4.4 (4.1) | 4.6 (3.8) | 0.772 |
| **Stroke classification** | PACS | 66 (65.3) | 35 (34.7) | 0.854 |
|  | LACS | 51 (60.7) | 33 (39.3) |  |
|  | POCS | 39 (66.1) | 20 (33.9) |  |
|  | TACS | 12 (54.5) | 10 (45.5) |  |
|  | Not recorded | 7 (63.6) | 4 (36.4) |  |
| **Emotionalism 6 months** | PSE | 16 (50.0) | 16 (50.0) | <0.001 |
|  | No PSE | 61 (48.0) | 66 (52.0) |  |
|  | Not recorded | 98 (83.1) | 20 (16.9) |  |

As is evident, there were no significant differences between participants whose carers completed MCSI compared to those who did not, on variables of sex, age, social deprivation, anxiety, depression and stroke classification. There was MCSI variation by TEARS recruiting centre, likely reflective of difference in resource rather than participant differences.

Furthermore, when PSE status was not recorded, MCSI data was also not recorded, largely explained by participants (and therefore carers) not attending for 6-month data collection.

As 175 participants did not return a measure of carer strain and only 102 did, we did not consider imputation methods for subsequent analyses. Instead, we continued noting that, based on participants characteristics, there was little evidence of any association with presence or absence of carer strain measurements, and thus no selection bias was suspected.

## Carer Strain

Scores on MCSI range from 0 to 26 and were distributed as per Figure 1. As is evident, the distribution was heavily skewed. For better modelling, we therefore computed sqrt.MCSI, plotted in Figure 2. Prior to transformation, the MCSI distribution had a median of 4.0 and a mean of 6.0. Following transformation, the distribution had a median of 2.0 and mean of 2.0.

**Figure 1:**
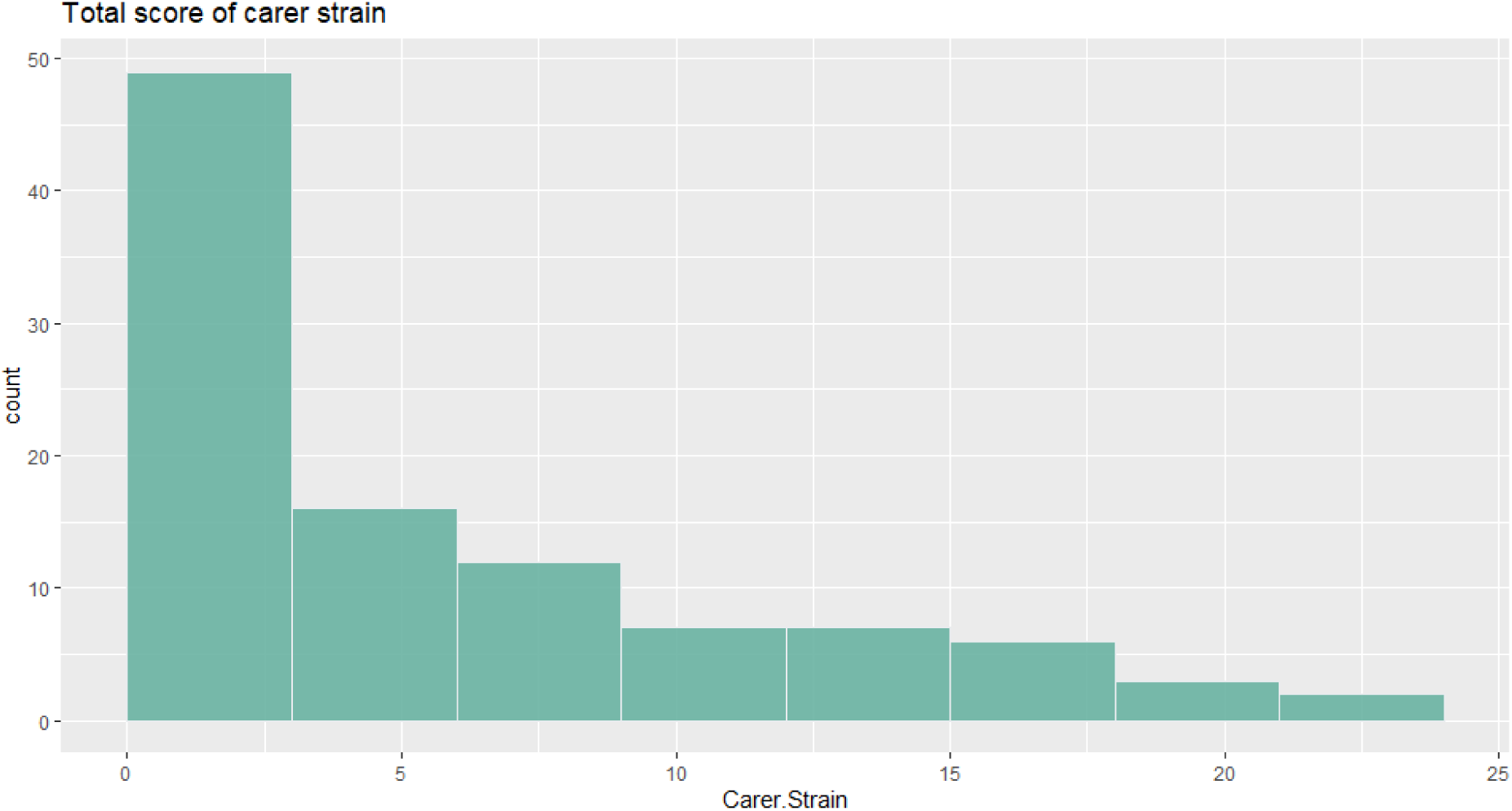
Histogram of MCSI total score.

**Figure 2:**
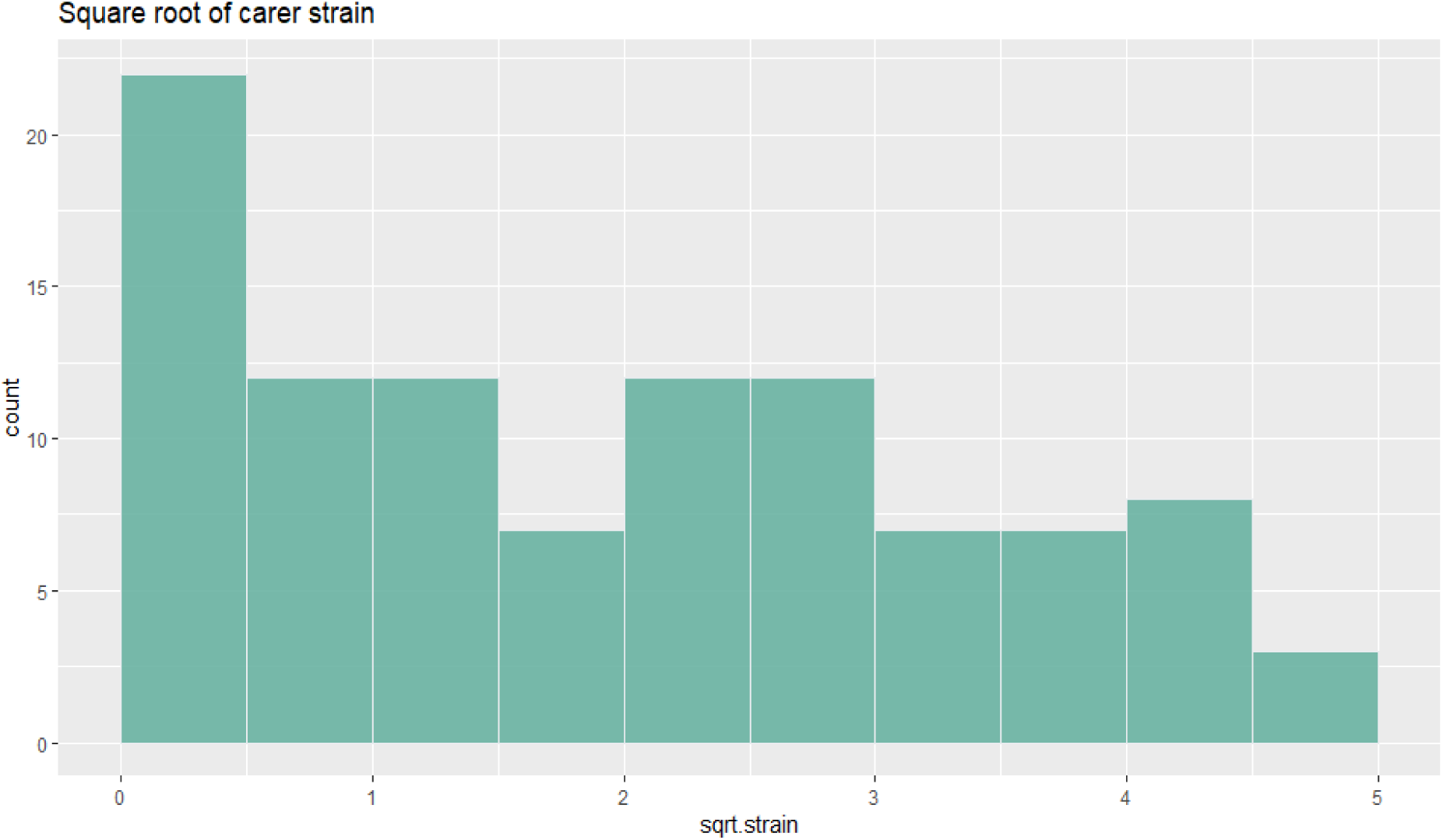
Histogram of square root transformed MCSI total score.

## Association of MCSI total score with appropriate putative predictors

Next we examined the association of participant sex, age, social deprivation, anxiety, depression, Barthel Index, stroke classification, carer strain and sqrt.MCSI by PSE status at 6-months.

As is evident in Table 2, anxiety was higher in patients known to have PSE, but not statistically significant in this sample. The Barthel Index was lower in PSE patients known to have PSE, suggesting PSE associates with greater functional dependence. The sqrt.MCSI stabilised the variance (and therefore the standard deviation), with carer strain higher for those with known PSE.

**Table 2:** Participant characteristics, cross-classified by PSE status at six months.

| **N of participants** | **PSE** | **No PSE** | **Not recorded** | **P** |
| --- | --- | --- | --- | --- |
|  | 16 | 66 | 20 |  |
| **Sex = male (%)** | 10 (62.5) | 37 (56.1) | 13 (65.0) | 0.736 |
| **Age at stroke (mean, SD)** | 56.75 (14.91) | 66.05 (13.06) | 71.55 (13.98) | 0.006 |
| **Social Deprivation (mean, SD)** | 2000.31 (1841.62) | 3341.26 (2093.65) | 2472.84 (1637.68) | 0.029 |
| **Anxiety (mean, SD)** | 7.33 (5.77) | 5.46 (4.47) | 3.62 (4.47) | 0.183 |
| **Depression (mean, SD)** | 4.40 (4.50) | 4.64 (3.63) | 4.62 (3.70) | 0.975 |
| **Barthel ADL (mean, SD)** | 17.88 (3.69) | 19.23 (1.58) | 14.74 (6.05) | <0.001 |
| **Stroke classification (%)** |  |  |  | 0.069 |
| PACS | 4 (25.0) | 23 (34.8) | 8 (40.0) |  |
| LACS | 6 (37.5) | 23 (34.8) | 4 (20.0) |  |
| POCS | 4 (25.0) | 14 (21.2) | 2 (10.0) |  |
| TACS | 1 (6.2) | 3 (4.5) | 6 (30.0) |  |
| Not recorded | 1 (6.2) | 3 (4.5) | 0 (0.0) |  |
| **Carer strain MCSI** | 9.75 (8.52) | 4.23 (4.89) | 8.75 (6.48) | <0.001 |
| **Sqrt MCSI** | 2.73 (1.56) | 1.61 (1.29) | 2.60 (1.44) | 0.001 |

To establish the associations between carer strain and patient characteristics, sqrt.MCSI was therefore regressed upon age, sex, deprivation (as measured by SIMD rank), Barthel Index, Stroke class (Oxford classification measured at baseline), and PSE status at 6 months.

Covariates that did not achieve statistical significance (by Wald test) were dropped to obtain a parsimonious final model. HADS anxiety and HADS depression were not included in this modelling since they were considered to arise from PSE and functional status (measured by Barthel Index).

The results of modelling are provided in Table 3 with univariable results included for reference, since these were used to suppress terms with little evidence of association to sqrt MCSI (P values > 0.1). As is evident, the final model had two covariates: Barthel Index and PSE status; these were the root sources of the associations within the data. Participants with a lower Barthel had higher associated values of sqrt.MCSI (carer strain). The size of the effect was such that a 6 unit decrease in Barthel Index, say from 20 to 14, associated with a unit (1.0) increase in sqrt.MCSI.

**Table 3:** Table of regression coefficients, with sqrt.MCSI as dependent variable.

|  |  | **Coefficient (univariable)** | **Coefficient (multivariable)** |
| --- | --- | --- | --- |
| **Sex** | Female | - | - |
|  | Male | 0.09 (-0.49 to 0.67, p = 0.756) | - |
| **Age at stroke** |  | -0.00 (-0.03 to 0.02, p = 0.630) | - |
| **Social deprivation** |  | -0.17 (-0.30 to -0.03, p = 0.19) | - |
| **Barthel ADL** |  | -0.14 (-0.21 to -0.06, p < 0.001) | -0.16 (-0.30 to -0.02, p = 0.026) |
| **Stroke Classification** | PACS | - | - |
|  | LACS | 0.09 (-0.60 to 0.77, p = 0.806) | - |
|  | POCS | 0.43 (-0.37 to 1.22, p = 0.291) | - |
|  | TACS | 1.14 (0.12 to 2.16, p = 0.029) | - |
|  | Unknown | -0.13 (-1.63 to 1.37, p = 0.868) | - |
| **PSE status 6 months** | No PSE | - |  |
|  | PSE | 1.13 (0.38 to 1.87, p = 0.004) | 0.95 (0.16 to 1.73, p = 0.019) |

Moreover, PSE was associated with a 0.91 point increase in sqrt.MCSI. Whilst it is difficult to practically assess patients on a sqrt.MCSI scale, the effect size of PSE on MCSI would be similar to a 6 point difference on Barthel Index.

Table 2 shows the unadjusted average carer strain of those with PSE to be 9.75 compared to an unadjusted average of 4.23 in those without PSE. Further modelling was undertaken to explore nonlinear effects of social deprivation, age, and Barthel Index. These are not reported since the effects were acceptable as linear.

Finally, we produced a violin plot with boxplot within, to reflects more details of the distribution of carer strain (Figure 3), including carer strain adjusted from the final model for the association with Barthel (Figure 4). As is evident, the distribution of MCSI data amongst carers of people with PSE has a higher median, greater inter-quartile range with more frequent low scores, which held for the distribution adjusted for Barthel.

**Figure 3:**
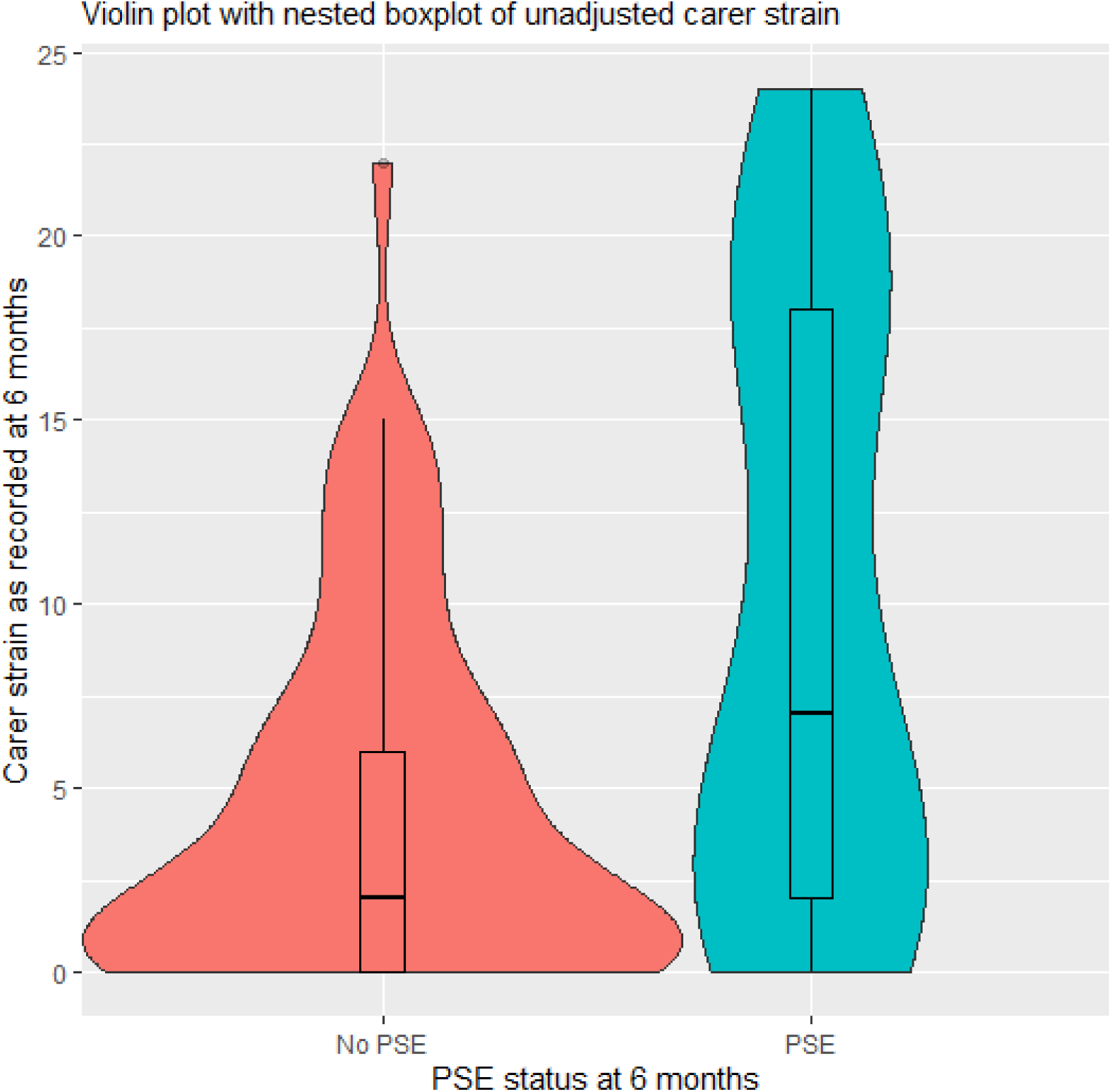
Violin plot with nested boxplot of unadjusted carer strain.

**Figure 4:**
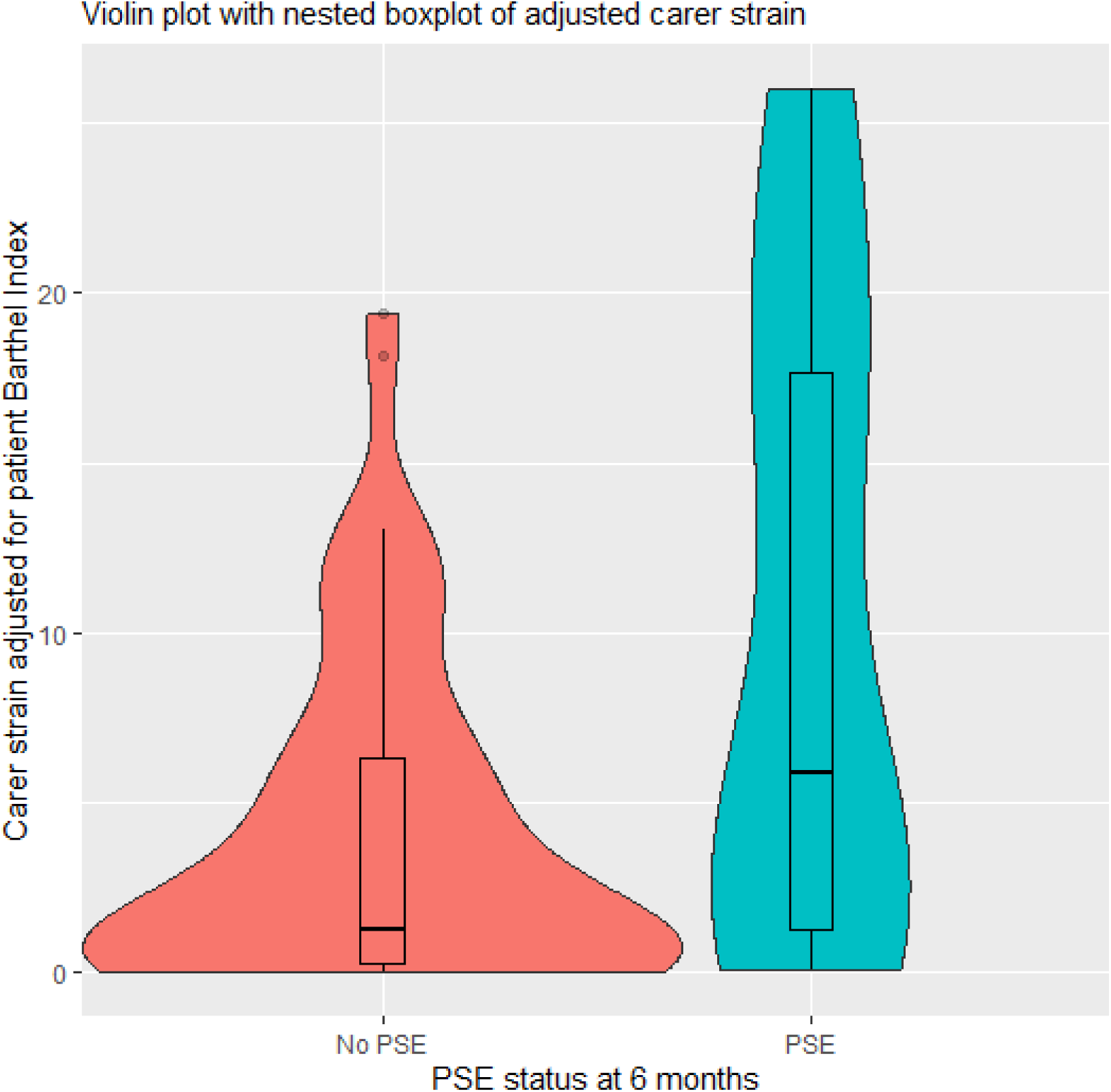
Violin plot with nested boxplot of adjusted carer strain.

## DISCUSSION

To the best of our knowledge, this is the first study to examine carer strain in the context of emotionalism after stroke. We deployed a widely used, validated measure of carer strain and compared the psychological impact of caring for people with stroke and emotionalism to the impact of caring for people with stroke but no emotionalism. We also considered whether carer strain varied with the particular characteristics of the cared for individual, including age, sex, stroke type, deprivation, mood and functional status.

Several key findings emerged. First, we did not observe any significant association between carer strain measured on MCSI and sex, age, deprivation level or stroke type of the cared for individual. By contrast, we observed a clear and substantial association between carer strain and functional dependence on Barthel Index, greater dependence associated with greater carer strain. Moreover, we also observed a significant association between carer strain and PSE status, strain being greater amongst carers of individuals with diagnosed PSE, compared to carers of individuals with stroke but no PSE. Consistent with these effects, when we regressed carer strain upon age, sex, deprivation, Barthel Index, Stroke type and PSE status at 6 months, the final model had two covariates: Barthel Index and PSE status. Again, higher carer strain was associated with greater functional dependence and diagnosed PSE.

Overall then, our analyses suggest a substantial association between functional dependence and carer strain but also between PSE status and carer strain, remaining after accounting for the impact of functional dependence measured by the Barthel Index. Consistent with the effects Calamonico and colleagues observed across the neurologic disorders, our analyses indicate caring in a PSE context significantly increases carer strain, equivalent to a six-point difference on the Barthel scale.

This is a unique study of carer strain in stroke patients assessing the impact of PSE on carers, the first to directly address this topic. We analysed MCSI data collected at 6-months post index event rather than acutely, so the findings offer a longitudinal analysis, albeit only at one time point. Our assessments were conducted face to face with participants and their caregivers rather than online. Colamonico and colleagues took an online survey approach, using the self-report Centre for Neurologic Studies-Lability Scale^27^ to denote emotionalism status of the cared for individuals. By contrast, we diagnosed PSE status using a semi-structured diagnostic interview, constructed based on House diagnostic criteria of PSE^1^, delivered face-to-face to people with stroke and their caregivers by stroke research nurses, pre-trained by the senior author, an emotionalism expert.

A number of study limitations must be highlighted. Although larger than the cohort of stroke caregivers surveyed by Colamonico, our sample size of 102 caregivers is still relatively small with data obtained from an observational study. Moreover, only 82 were associated to stroke survivors with known PSE status and with MCSI data returned, and only sixteen were carers of people with PSE. Replication will be required using a larger cohort of stroke survivors with PSE and their carers, although our sample is similarly modest to that of Colamonico and colleagues (59 stroke caregivers) who observed a similar effect.

The measurements were carried out as part of the follow up of the TEARS cohort study and thus represent observational data. Further, the data were only collected at a 6-months follow up session whenever the carer was present with the patient and willing to participate. Our conclusions must be limited as they may not apply to where the patients have more severe stroke (see Table 1 for details). Consequently, they should be regarded as a convenience sample at one time point. Nonetheless there is no suspicion of bias in the findings and participation of carers appeared to be representative of the observed characteristics of the patient population. Finally, whilst MCSI has acceptable psychometrics^14^ and previous use in stroke^19^ the scale lacks established cut points. Whilst this makes it hard to determine the clinical extent of carer strain seen in our data, the unadjusted average carer strain of those with PSE is 9.75 compared to 4.23 in those without PSE. The additional impact of caring in the PSE specific context is thus clearly evident.

### Clinical Implications

Whilst from a preliminary observational study and only at one time point, the data have important clinical and research implications. Clinically, additional psychological strain might be expected to arise in carers of individuals with PSE, something stroke clinicians and rehabilitation teams should hold in mind when PSE is detected. Targeted screening of carers of those with emotionalism using MCSI and other reliable measures could form an additional element of stroke care. Strain amongst carers of those with chronic PSE might be expected to escalate beyond 6-months, although larger scale longitudinal research beyond 6-months will be required to determine this. Research using qualitative approaches is needed to improve our understanding of the lived experience of caring for someone with emotionalism following stroke, including what can help both the patient and the carer, psychologically. This important research could in turn inform badly needed work to adapt current evidence based psychological interventions aimed at reducing carer strain in the broader stroke context and then test these for the PSE context.

## Data Availability

original data available from the corresponding author, although for data protection an embargo period may apply.

## Declaration of conflicting interests

None

## Acknowledgements

We sincerely thank our stroke nurse, SSRN and NHS GGC R&D colleagues.

## Funding

Stroke Association, UK.

## Notes

### Competing Interest Statement

The authors have declared no competing interest.

### Funding Statement

This research was funded by the Stroke Association UK (TSA 2013/03)

### Author Declarations

This study was approved by Scotland AREC Ethics Committee ID 157483

